# COVID AMP: An Open Access Dataset of COVID-19 Response Policies

**DOI:** 10.1101/2023.05.01.23289163

**Authors:** Rebecca Katz, Kate Toole, Hailey Robertson, Alaina Case, Justin Kerr, Siobhan Robinson-Marshall, Jordan Schermerhorn, Sarah Orsborn, Michael Van Maele, Ryan Zimmerman, Tess Stevens, COVID AMP Coding Team, Alexandra Phelan, Colin Carlson, Ellie Graeden

**Author notes:** corresponding author(s): Ellie Graeden. A full list of members appears in the Author Contributions (COVID AMP Coding Team).

## Abstract

As the COVID-19 pandemic unfolded in the spring of 2020, governments around the world began to implement policies to mitigate and manage the outbreak. Significant research efforts were deployed to track and analyse these policies in real-time to better inform the response. While much of the policy analysis focused narrowly on social distancing measures designed to slow the spread of disease, here, we present a dataset focused on capturing the breadth of policy types implemented by jurisdictions globally across the whole-of-government. COVID Analysis and Mapping of Policies (COVID AMP) includes nearly 50,000 policy measures from 150 countries, 124 intermediate areas, and 235 local areas between January 2020 and June 2022. With up to 40 structured and unstructured characteristics encoded per policy, as well as the original source and policy text, this dataset provides a uniquely broad capture of the governance strategies for pandemic response, serving as a critical data source for future work in legal epidemiology and political science.

## Background & Summary

In response to the COVID-19 pandemic, governments around the world implemented a range of policies, regulations, and mandates to mitigate transmission, support the economy, and protect population health. Despite targeting similar goals, there was significant heterogeneity in how governments approached policy strategies for the pandemic response, in part because of a dearth of prior policy evidence, an evolving understanding of which governmental actions might effectively protect populations, differing access to resources required for specific policy actions, and mismatched expectations regarding adherence to stringent policies.

Most policy trackers deployed during the pandemic focused on social distancing measures with an emphasis on the ability to assess the effectiveness of these policies in limiting human movement, human-human interaction, and disease spread as quantified by reported cases, hospitalizations, and fatalities for different populations and subpopulations. [1] These efforts, while critical in performing data capture for rapid analysis of the relative value of different social distancing measures, did not capture the full breadth of policy measures implemented, limiting policymakers’ ability to assess the impact of these “non-health” policies and the synergistic effects of a more integrated approach to pandemic response. [2]

To address this gap, the COVID Analysis and Mapping of Policies (COVID AMP) dataset tracked policy responses to COVID-19 around the world between January 2020 and June 2022. [3] We focused on high-volume data collection in real-time during the event, which allowed us to document and retain a historical record of policy changes, even as policy and guidance was being posted and its online record replaced, risking erasure. Data collection prioritized breadth over depth, while including comprehensive data collection for a subset of specific regions and topics to facilitate comparative analysis. Therefore, in some cases only a single or a few policies were captured for jurisdictions outside of the United States.

The COVID AMP dataset archives official legal documents and/or policy announcements at the local, intermediate, and national levels, including details such as the dates they were active, tags related to public health and economic relief, and up to 40 other characteristics per policy.

[3] The coded fields are aligned with the key sectors defined during prior outbreaks and established by existing emergency management frameworks, and the breadth of the data supports cross-sector and combinatorial analysis of policy impact.

With nearly 50,000 policy measures, regulations, and announcements, COVID AMP provides a new and powerful foundation for secondary analysis to better assess the impact of all policies implemented to manage the pandemic response. Taken together, these data are a critical addition to the body of work describing the policy and governance response to COVID-19 and a significant advancement in how we understand the heterogeneity of policymaking during outbreak response.

## Methods

### Data Collection

Beginning in April 2020, we identified policies implemented to mitigate and respond to the COVID-19 outbreak from January 2020 through June 2022. Policies were defined broadly and included the actions described and/or authorized in signed legislation, executive orders, ministry regulations, official press releases, and social media announcements made by verified authorities. We reviewed official government websites, databases, and social media pages to source documents (e.g., public health ministry websites, legislative archives, published press releases) for each jurisdiction.

For data entry, we designed an Airtable base (https://airtable.com) for researchers to collect and code policies. Each coder was trained by a lead researcher on how to navigate Airtable, source policies, and code each policy. Before inputting records to the official dataset, researchers were required to code a standard set of practice policies to assess competence and inter-coder reliability. Once they met these criteria, coders were moved to the main Airtable base. Each researcher received a specific jurisdiction assignment, as each jurisdiction publicized and released policy information to the public differently. Data collection efforts initially prioritized capture of policies implemented in the United States, but as coverage was extended globally, researchers with language skills or lived experience in a country were given priority assignment to those jurisdictions. Researchers met on a weekly basis to discuss emergent themes, new category/subcategory/target types, and answer questions about coding processes or definitions. The lead researcher also held office hours to assist with policy sourcing and coding, as well as to perform regular review and technical validation of researchers’ progress.

To source policy data for the United States (U.S.) and its territories, we reviewed websites for the state governor’s office, Department of Health or comparable agency, and if applicable, the state legislature. Researchers consulted the POLITICO Pro Legislative Compass to identify additional state-level COVID-19 legislation. [4] For other countries, researchers identified the primary authorities for health policy and reviewed their official websites for policies. In addition, researchers used search engines for COVID-19 policies in the local language. If the coder was not fluent in the local language of the jurisdiction being collected, Google Translate (https://translate.google.com) was used to translate policy documents. The Internet Archive was used to identify policies in circumstances where policies were removed or updated from the original site. [5] Only official government policies issued in direct response to the COVID-19 pandemic were coded in the dataset. In cases where an official record of the policy could be found but where the government document(s) for the policy were not published, no longer available, or inaccessible from the Internet Archive, we coded the record from social media records, news notices, and policy announcements describing the official government policy.

### Data Structure & Coding Process

As we collected polices, we concurrently and iteratively developed a coding scheme to capture structured and unstructured data related to COVID-19 response policies, balancing an internally-consistent taxonomy with flexibility to describe heterogenous policy environments. Many policies include one-to-many relationships in which a single policy established more than one and often several different directives related to COVID-19 mitigation or response management. Each row in the dataset represents an individual directive, linked by a unique identifier to the original policy document and coded by the type of policy and the target of policy, as defined as the primary population, location, or entities impacted by the policy or law, in addition to more than 40 additional coded observations per directive. [3]

Event response and event-specific mitigation efforts are only one subset of the policies needed to effectively manage and respond to large scale emergencies. [6] The National Response Framework (NRF) in the U.S. lists 12 emergency support functions and leans on a whole-of-government response framework to manage critical functions across transportation, military authorities, manufacturing, supply-chain management, first-responder housing, cross-border licensing issues for critical response personnel (e.g., nurses, electrical lineman), housing authorities, and economic support for those impacted. [7] Building on this cross-sector approach, previously identified and applied in the Georgetown Outbreak Activity Library (https://outbreaklibrary.org/), we identified five categories of policy relevant to the COVID-19 outbreak: (1) Social distancing, (2) Emergency declarations, (3) Travel restrictions, (4) Enabling and relief measures, and (5) Support for public health and clinical capacity. Over the course of data collection, five additional categories emerged from policy analysis that were added to the coding scheme to more accurately capture the range of policy actions available and pull forward specific types of policies as they gained global traction (e.g., variation in vaccination policies): (6) Face mask, (7) Contact tracing and testing, (8) Military mobilization, (9) Authorization and enforcement, and (10) Vaccinations. In addition, 71 subcategories were used to capture the type of policy actions at a more granular level. For example, the social distancing category is composed of subcategories such as “Curfews”, “Event delays or cancellations”, “Alternative election measures”, “Private sector closures”, or “Stay at home.”

As the pandemic unfolded, the policies implemented by governments to manage the response and mitigate impacts evolved. Therefore, categories, subcategories, and targets were adjusted over time to maximize the taxonomy of the dataset for exhaustiveness and usability for secondary analysis. All updates to categories or subcategories were made by consensus of the research team, and backpropagated across existing data to ensure internal consistency. For a full Data Dictionary, see Supplementary Table 1.

### Comparison to other COVID-19 policy trackers

The COVID-19 pandemic prompted over 200 research and government initiatives aimed at tracking the policies and measures implemented in response to the outbreak. [1] Given the extensive nature of these efforts, a comprehensive evaluation of each one is beyond the scope of this article. However, we summarize the key features of COVID AMP [3] in the context of similar datasets, including OxCGRT [8], CoronaNet [9], and State Policy Responses to COVID-19 (SPRC19) [10] to highlight differences and provide suggestion for what types of subsequent analysis might be most useful.

The primary goal of COVID AMP was to capture a broad representative sample of the types of policies implemented to respond to a global pandemic. CoronaNet shares a similarly broad scope, identifying 20 “broad policy types” including NPIs, declarations of emergency, travel restrictions, health communication, and some public and private restrictions. [9] OxCGRT includes 19 indicators focused more specifically on containment, health, and economic support policies [8]. In the COVID AMP ontology, these “broad policy types” and “indicators” are equivalent to our concept of “Policy subcategory”, which includes more than 70 individual policy subtypes. [3] While COVID AMP encompasses many of the policy types included in other datasets, it is unique in the granularity captured, particularly with regard to economic policy measures. The dataset captures specific economic interventions, from tax delays to stimulus payments and anti-price gouging measures. [3]

In contrast to OxCGRT, we do not assign quantitative values to interpret policy stringency, but instead classify policies as either restricting or relaxing based on the intended effect of the directive on the policy environment at the time of enactment. This is a marked divergence from other policy trackers; this approach supports the ability to analyse not only patterns of lockdown, but the progressive reopening across jurisdictions through extensions, amendments, expirations, and repeals of policy [8,9].

To better understand the context of the polices implemented, the COVID AMP dataset includes details about the sector and demographic targets. In OxCGRT, these variables are binary for each of the indicators to simply designate “targeted” or “general.” [8] In CoronaNet, the demographic target aligns with 11 broad demographic targets or 25 special demographic targets; sector targets are not included. [9] Similar to CoronaNet’s fields “init_country_level” and “geog_target_level”, COVID AMP uses the terms ‘authorizing areas’ and ‘affected areas’ to define conditions in which a policy issued by one level of government applies to another geography. [3] For example, the United Kingdom’s travel restriction after the Omicron variant was identified in December 2021 targeted six countries with the United Kingdom designated as the “Authorizing country” and South Africa, Botswana, Lesotho, Eswatini, Zimbabwe, and Namibia each listed in the “Affected country” field. [11]

Beyond the geographic targets, COVID AMP uses the field “Policy target”, with 73 multi-select options to indicate the populations, places, sectors, or entities affected by the policy. In this way, researchers can mix and match specific policy categories/subcategories with sector and demographic targets to filter and conduct highly customized analysis. For example, which states required testing in primary secondary schools versus those that required tests only for higher education? What types of policies were used to prevent transmission of disease in farming, agricultural, and food processing facilities? Where were pharmacies allowed to dispense emergency medication refills? While the policy targets allow substantial flexibility compared to other resources, not all place types could be captured individually within the scope of COVID AMP. Those needing precise indicators about particular demographics, places, or sectors (e.g. only interested in beaches within the “Outdoor recreation/campgrounds/beaches/parks” target) may need to conduct additional review to extract the data most relevant to their research needs.

Because the focus of COVID AMP was to emphasize breadth of policy types, OxCGRT and CoronaNet have more extensive global coverage (184 and 195 countries, respectively). Although data from 150 countries is captured in COVID AMP, only 40 of these countries have more than 100 policies coded (see Supplementary Table 2). Thus, given the absence of comprehensive global coverage, it is recommended that COVID AMP data is triangulated between additional data sources for policies of interest when conducting cross-sectional analysis between countries.

For U.S. policies, COVID AMP offers a rich source of data through June 2022 at the state level, and through December 2021 at the county and tribal levels. Compared to similar U.S.-focused datasets like the SPRC19 dataset [10], the inclusion of county and tribal policies appears to be unique to COVID AMP. The COVID AMP dataset provides similar levels of policy detail as the SPRC19 dataset at the U.S. state level, but its temporal coverage enables longitudinal analysis of the policy strategies implemented by U.S. states, whereas SPRC19 currently only covers January to April 2020. [3, 10]

## Data Records

The COVID AMP library contains more than 15,000 individual COVID-19-related documents issued or effective through the period January 2020 to June 2022 (and beyond for select jurisdictions). A static version of the dataset, including the Data Dictionary and all raw PDF files, has been deposited in Zenodo [3]. Figure 1 shows the extent of policy data coverage globally and in the U.S.

**Fig. 1.**
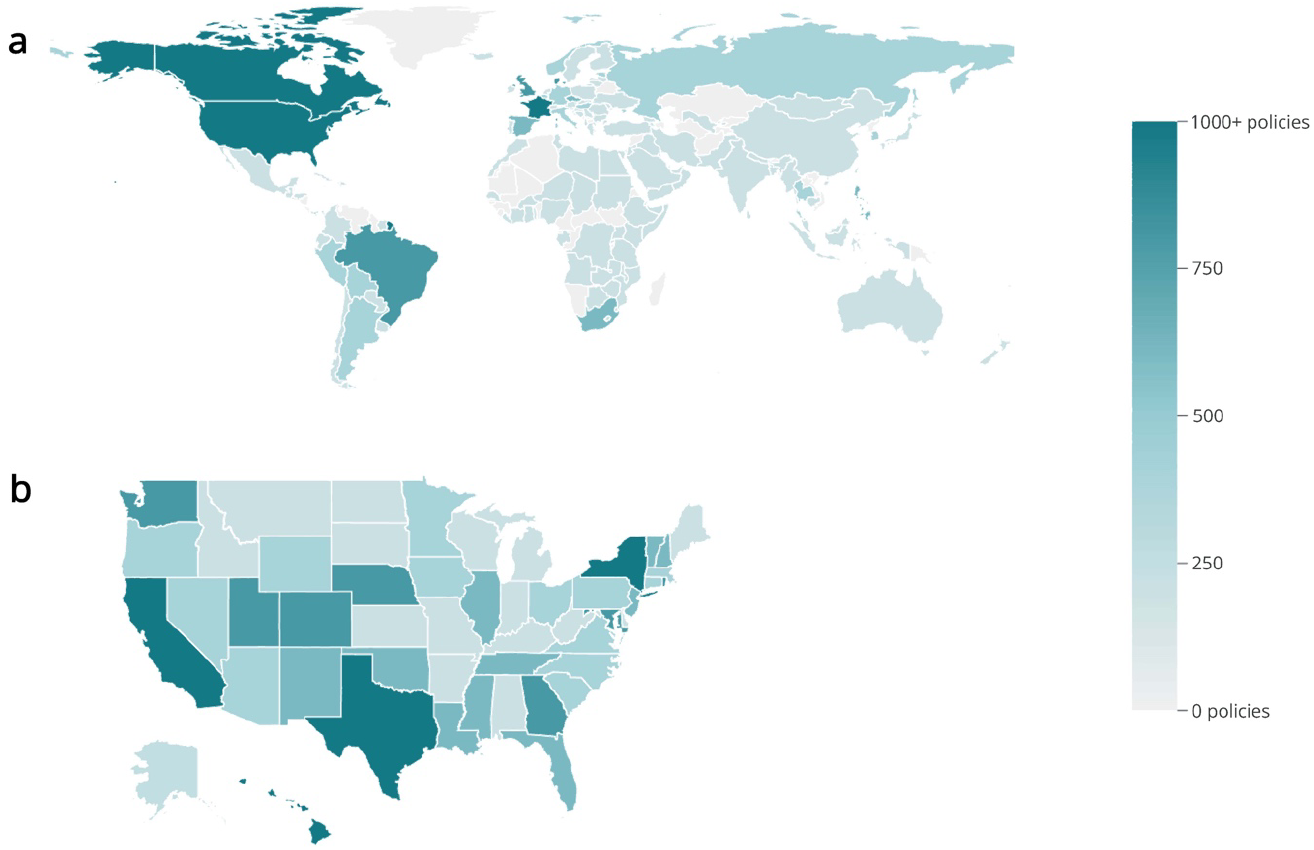
Geographic policy data coverage of the COVID-AMP dataset from January 2020 to June 2022 (a) Total number of policies captured for each country. There are 152 countries for which at least 1 policy is coded. (b) Total number of policies captured for each U.S. state. All 50 states and territories were coded comprehensively.

This dataset was designed to capture the breadth of measures applied by different jurisdictions to manage and mitigate the pandemic. Each individual measure included in a within a policy document with a unique policy target and subcategory is treated as a single policy with its own row. Therefore, a single policy document may be represented by many rows in the dataset. For example, an executive order can include a stay-at-home order for individuals and mandate non-essential business closures. While these policy directives share a common policy name and PDF as part of the same larger piece of legislation, they are captured as distinct entries (“policies”) with unique IDs. Extensions of previous policies are also captured as a new row and linked back to the previous policy using unique IDs.

**Table 1.** Required data fields and definitions

| **Data field** | **Field definition** |
| --- | --- |
| Unique ID | A unique identifier associated with data in each row. The data is captured so that each row represents a single policy action, per date issued, per authority, and per area affected. |
| Attachment for policy | PDF or image of the policy (permanently hosted in Amazon S3 bucket) |
| Data source for law/policy | URL from which to access the underlying law or policy, may be from the Internet Archive [5] |
| Policy name | The complete title of the law or policy, including any relevant numerical information |
| Policy number | A numeric identifier given to each policy release, including capturing co-released policies where applicable. A single policy release may contain multiple directives |
| Policy description | A written description of the policy and the directive by the researcher |
| Policy type | The type of policy that is enacted (e.g., executive order, emergency declaration, statute, etc.) |
| Policy relaxing or restricting | Broad designations about a policy with regard to its intended impact on the policy environment at the time the policy was issued. |
| Policy category | Categorization of the overall scope of the policy directive (e.g., social distancing, emergency declarations, travel restrictions, enabling and relief measures, support for public health and clinical capacity, contact tracing/testing, military mobilization, face masks) |
| Policy subcategory | Detailed information about the intention of the policy (e.g., face coverings, quarantine, private sector closures, school closures, etc.) |
| Policy target | The primary population, location, sector, or entities impacted by the policy or law (e.g., restaurants/bars, nursing homes and/or assisted living, essential workers, suspected cases, etc.) |
| Authorizing level of government | The level of government that authorized and/or issued the policy (e.g., global entity, country, intermediate area, local area, tribal nation) |
| Authorizing body | The office of the authorizing entity who issued the policy (e.g., governor, mayor, health official, president, city council, etc.) |
| Authorizing country name | The name of the country in which the authorizing entity is located |
| Affected country name | The name of the country to which the policy applies, if different from the country from which the policy was enacted |
| Issued date | The date on which the policy was initially announced and/or issued |
| Effective start date | Date on which the policy took effect or was enacted |
| Anticipated end date | The date on which the directive specified in the policy was intended to end |
| Actual end date | The data on which the directive specified in the policy was terminated, replaced, or extended |

Each policy directive is tagged with a series of descriptive attributes based on a detailed review of the policy language, including the following selection of fields:

A Data Dictionary describing each of the fields is available in Supplementary Table 1.

Figure 2 shows the distribution of policies by category over time, globally and within the U.S. Both globally and in the U.S., the greatest number of policies were initiated in April 2020; “Social distancing” was the predominant category of policies enacted by governments over the course of the pandemic, followed by “Enabling and relief measures” and “Support for public health and clinical capacity.”

**Fig. 2.**
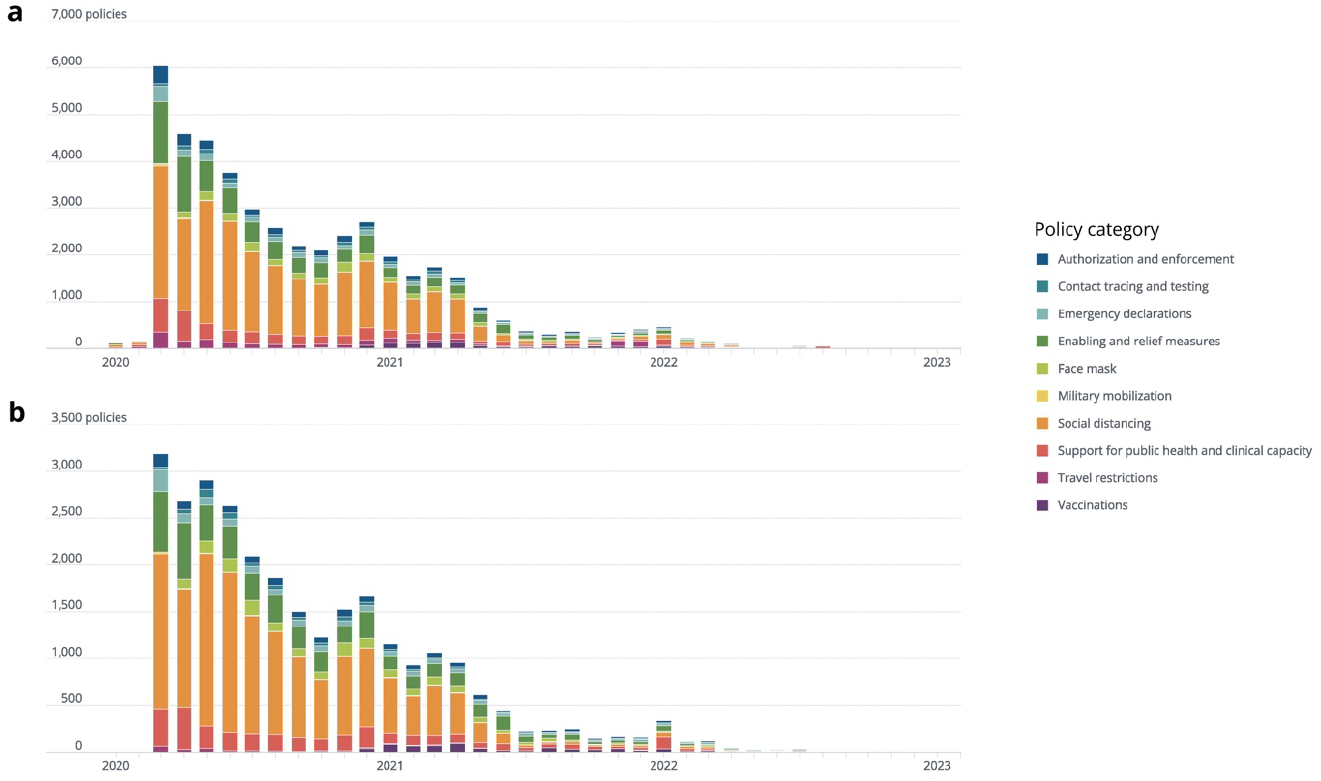
**(a)** Distribution of policies by category and month, globally. **(b)** Distribution of policies by category and month for the United States. The month for each policy was the effective start date.

In conducting analysis, users should expect to see a broad sample of the heterogenous policies implemented over the course of the COVID-19 pandemic, but COVID AMP is not a complete global historical record nor is it intended to be a comprehensive or complete description of all policies implemented globally. That said, to the best of our knowledge, the dataset contains a comprehensive set of policies implemented for each of the U.S. states, Puerto Rico, and Guam from January 2020 to June 2022, with over 20,000 policies captured at the national and state level in these jurisdictions. [3] Among U.S. state and national policies, 95% were issued prior to November 23, 2021. This decline in the number of policies issued corresponds with the desire of many U.S. states to “return to normal” by early 2022. Researchers also documented many policies for U.S. counties (approximately 8,000 policies) and tribal areas (approximately 1,400 policies), however, comprehensive coverage is limited to California, Washington D.C., Maryland, Nevada, and Virginia from January to December 2020 by request to support the pandemic response in these states. [3]

For jurisdictions outside the U.S. and its territories, policy coverage is generally lower, with researchers focusing data collection efforts at the national level as opposed to the intermediate area levels (e.g., state, province, etc.). These decisions were made largely ad hoc and based on language skills on the research team. Therefore, the distribution of policy counts across countries varies significantly. Out of the 150 countries included in the dataset, 40 have more than 100 policies coded, while 77 countries have 5 or fewer policies recorded (see Supplementary Table 2). The limited number of recorded policies (between one and five) for certain countries over a discrete period (November and December 2021) is attributed to a specific data collection effort focused on assessing travel bans related to the Omicron variant in late fall of 2021. [11]

Due to variations in legal systems, the significance of the total number of policies varies by state and country. Some jurisdictions regularly renewed the emergency authority of health departments or other bodies, thus re-issuing the same policies regularly and appearing to have more policies by total number. Thus, policy totals tend to reflect variation in governance structure and method more than stringency or effectiveness of the policy response.

## Technical Validation

Given the frequency and scale of data collection, the research team implemented a combination of manual and automated quality assurance and control (QA/QC) processes to check and correct the data. The manual QA/QC process involved a lead researcher who reviewed data for typographical errors, ensured inter-coder reliability, and confirmed record completion. Completed records included all fields specified in the “Data Records” section; records with missing fields were flagged for review and excluded until corrected. The fields, “Anticipated end date” and “Actual end date” were exceptions, as many policies did not specify the intended end date or were ongoing during data collection. Policies for which the end date is not provided may still be in place or permanent or the end date for the policy was not publicly documented. Approximately 50% of policies have an “Anticipated end date” and 70% have an “Actual end date.”

In addition to manual review, automated QA/QC was applied to clean and standardize the data. Drop-down lists, with easily accessible data definitions and glossaries, were used to standardize coding selections, prevent typos, and reduce discrepancies. The Dedupe extension in Airtable (https://airtable.com) was used to find and manage duplicate records based on policies with identical issued/effective start dates, authorizing/affected areas, and data sources. Where a duplicate was identified, the lead researcher merged the information from the two records, selecting the correct information from each if discrepancies in coding existed. Python (version 3.7.0, https://python.org) was used to filter incomplete records out of the final dataset view, assign policy numbers, and standardly format dates and country names for ease of use in secondary analysis.

## Usage Notes

From the implementation of mask mandates to the reduction of prison populations and alternate measures of voting during elections, the COVID AMP library contains a wide array of policy documents that historians, legal experts, economists, and epidemiologists can analyze to assess and compare the effectiveness of COVID-19 outbreak responses around the world. Pairing this tool with epidemiological data supports the evaluation of policy effectiveness and how that success relates to the affected population, authorizing entity, health infrastructure, and other extenuating factors. We also hope that this library will support policymakers in future outbreaks by providing canonical examples of policies from countries and states that had different outcomes during the pandemic.

### Example Analysis

The data collected in COVID AMP provides researchers with valuable insights to understand how policy is used to respond to a global pandemic and inform policy response for the next. The granularity of the COVID AMP dataset allows for disaggregation and analysis by subcategory of policy enacted. Figure 3 shows one example of the type of analysis supported by this approach and shows the diversity of economic policy measures implemented over time for the U.S. From February 2020 to February 2022.

**Fig. 3.**
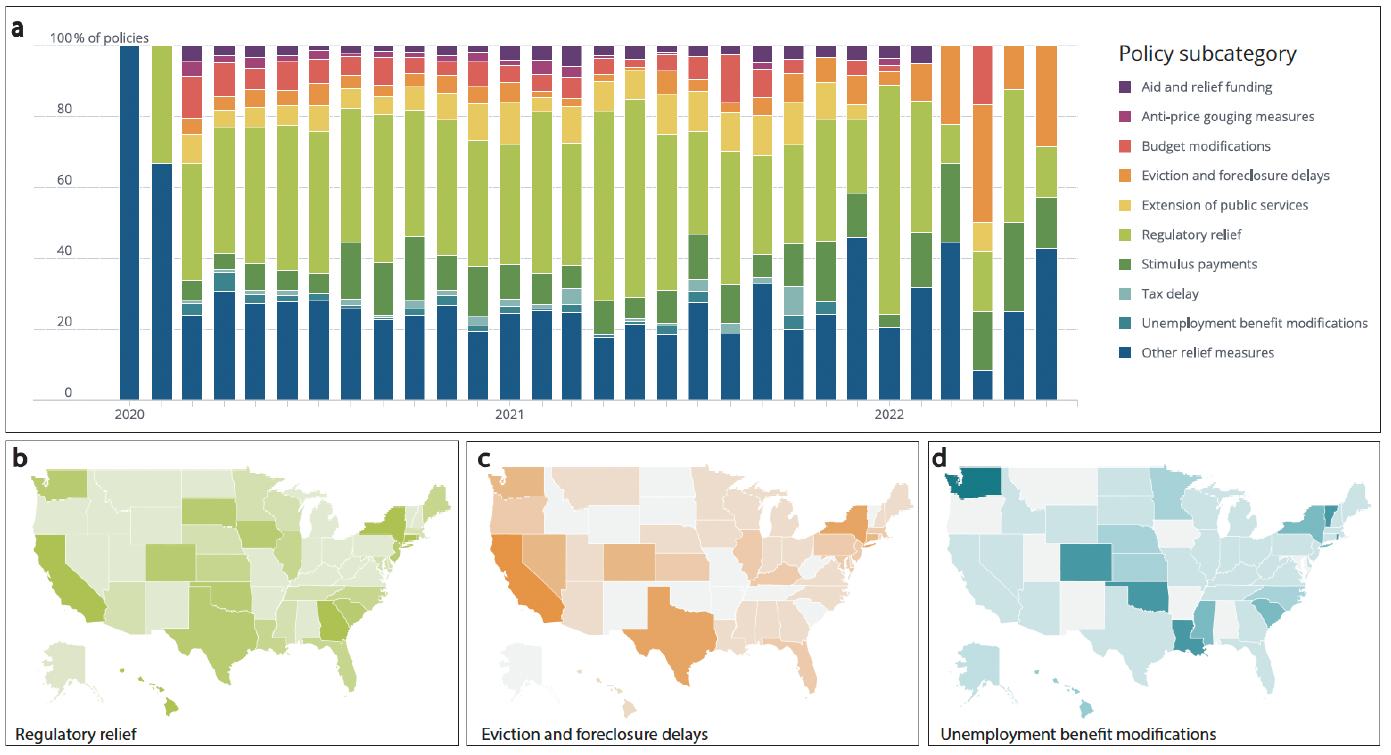
Economic policies implemented in the United States as of June 2022. **(a)** Relative proportion of economic relief policies (from the “Enabling and relief category) enacted over time by policy subcategory. **(b)** Relative number of regulatory relief policies implemented at the state-level. **(c)** Relative number of eviction and foreclosure delay policies implemented at the state-level. **(d)** Relative number of policies implementing modifications to unemployment benefits at the state-level.

As shown in Figure 3a, regulatory relief made up the largest proportion of economic measures implemented in the United States for the majority of the pandemic and was only overtaken by eviction and foreclosure delays as of early 2022. The widespread adoption of regulatory relief measures can be seen in Figure 3b. In addition to these national-level trends, individual-level economic support policies varied widely across the United States reflecting the different strategies employed by different states. For example, states including New York, Texas, and California implemented eviction and foreclosure delay policies as shown in Figure 3c. By contrast, Washington, Colorado, Oklahoma, and Louisiana implemented comparatively more modifications to unemployment benefits to provide support to those affected by the pandemic (see Figure 3c). A comprehensive analysis of these strategies, combined with other key economic indicators, offers valuable insights to researchers studying the unique characteristics of the population and economies of different states and the impact of these types of economic policies in mitigating the pandemic and/or reducing economic harms.

COVID AMP also supports analysis of the intended targets of each policy implemented, as shown in Figure 4. For example, economists could use the data to assess specific policy types, such as “Regulatory relief”, to analyse which sectors received which types of support and compare effects across jurisdictions and sectors. Using the date each policy was issued and became effective, analysts could, for example, assess how stock prices reacted to regulatory relief announcements. For education officials, the COVID AMP data could be combined with school test scores to understand how the timing of school closures, reopening, and distance learning impacted students’ educational performance. Public health researchers could use the data to identify a specific population, such as “Homeless shelters and individuals” and determine which policy types were (or were not) targeted toward the population, and whether it met community needs. With the ability to cross-reference policy subcategories and targets, COVD AMP enables researchers from various fields to conduct more nuanced analyses of the impact of policies on their area of interest, whether that is a sector, population, or policy type, and encourages policy innovation for the future.

**Fig. 4.**
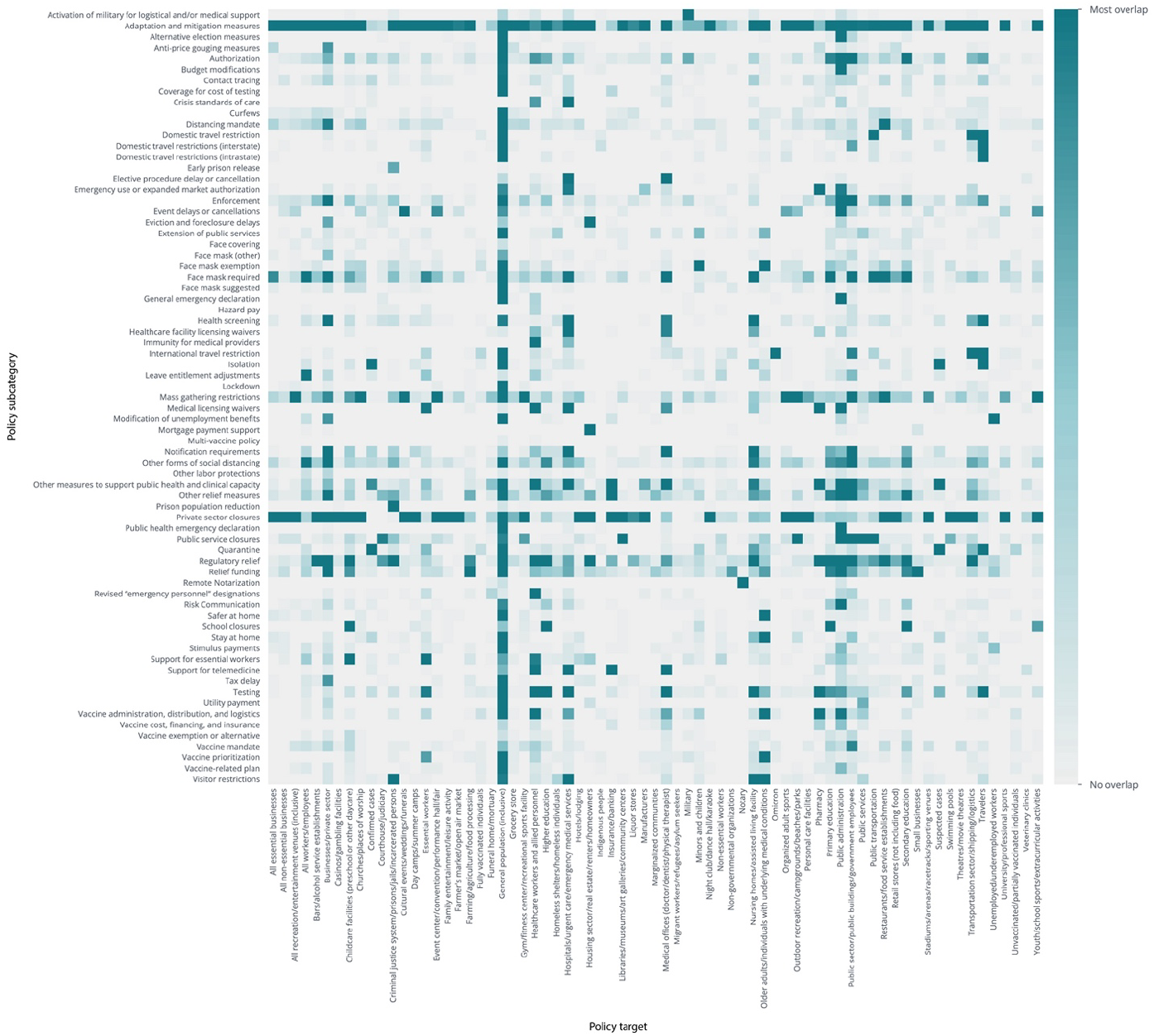
Heat map shows the co-occurrence of policy subcategories and policy targets globally, with the darkest squares representing the most overlap. The x-axis shows lists the policy targets, and the y-axis lists the policy subcategories.

### Published Research Using COVID AMP

As a library of policies collected in near real-time and continuously evolving throughout the pandemic, COVID AMP allows users to identify and access policies of interest in addition to the original text of the policy as a raw text file or PDF [3]. These data can then be used to perform secondary data transformation as needed for derivative analysis. The COVID AMP dataset does not prescribe research-side assumptions such as policy stringency or policy levels to the data with the specific intent of supporting broader cross-disciplinary downstream use. The value of this approach is demonstrated by Page-Tan & Corbin (2021), who used COVID AMP data to define unique parameters of restrictiveness to test four different policy scenarios in states and localities with high social vulnerability scores using propensity score matching. [14] Additional studies used COVID AMP to validate parameter assumptions for models about the timing of intervention implementation [15, 16]. Others have used COVID AMP to analyse global differences in response strategies to the Omicron variant through specific focus on travel restrictions [11], access archived public health measures from governments, trace the progression of policy, evaluate the role of institutions [17, 18], and assess the benefits of mask-wearing at the county-level. [19] These studies highlight the ease of use of the dataset and suggest that significant future work could continue to make use of COVID AMP to ask new questions about the COVID-19 pandemic.

## Data Availability

All data produced are available online at Zenodo DOI: 10.5281/zenodo.8087600

https://zenodo.org/record/8087600

https://covidamp.org/

## Code Availability

COVID AMP data are available via an application programming interface (API) and are licensed under the Creative Commons Attribution CC BY Standard at: https://api.covidamp.org/docs.

We provide a public, interactive web interface for visual exploration of the dataset at: https://covidamp.org/. Within the site, data is available at: https://covidamp.org/data?type=policy. This page allows for download of the full dataset or filtered subsets of the data. Additionally, documentation of the methods, including a data dictionary and glossary, are available at: https://covidamp.org/about/doc.

In addition to this manuscript, a static version of the COVID AMP dataset itself can be cited directly as Zenodo DOI: <u>10.5281/zenodo.8087600</u>

All policies and directives coded within the dataset have been reviewed and technically validated. We hope that the dataset will support research efforts aimed at improving pandemic response strategies and inform future outbreak policy analysis.

## Acknowledgements

Funding for COVID AMP provided by Rockefeller Foundation, NTI Bio, and Georgetown University.

## Author contributions

Rebecca Katz conceived of the idea, designed and managed data collection, provided subject matter expertise and guidance, and reviewed the manuscript.

Ellie Graeden conceived of the idea, designed and managed data collection, provided subject matter expertise, led design and build of the data platform, and drafted the manuscript.

Kate Toole helped draft the manuscript, managed data entry, trained coders, performed QA/QC, and performed analysis.

Hailey Robertson helped draft the manuscript, performed QA/QC, prepared the dataset for publication, performed analysis, and produced visualizations.

Alaina Case managed data entry, trained coders, contributed to design of the data platform, user interface, and web access for the data.

Justin Kerr contributed to design and build of the data platform, user interface, and web access for the data, and served as subject matter expert on data structure.

Siobhan Robinson-Marshall managed data entry, trained coders, and performed QA/QC. Jordan Schermerhorn managed data entry, trained coders, and performed QA/QC. Sarah Orsborn managed data entry, trained coders, and performed QA/QC.

Michael Van Maele contributed to design and built the online data platform, user interface, and web access for the data.

Ryan Zimmerman contributed to design and built the online data platform.

Tess Stevens designed the data platform, user interface, and web access for the data. Alexandra Phelan contributed to project conception and provided legal guidance.

Colin Carlson provided input on data visualization.

The **COVID AMP Coding Team** identified policy sources, reviewed and coded policies, and performed QA/QC. Team members include:

Yasser Omar Abdellatif, Omolara Adekunle, Saba Alfred, Madison Alvarez, Ariyand Aminpour, Jennifer Ayres, Alice Bolandhemat, Matthew Boyce, Anjali Britto, Josephine Bryar, Sophia Byrne, Andrea Cano, Ethan Cantrell, Tianhui Cao, Yujie Chen, Kahiau Cockett-Nagamine, Kayleigh Coppinger, Katie Dammer, Julia Damski, Nathalie Danso, Aleena Dawer, Rose Dever, Maydha Dhanuka, Roma Dhingra, Maria Victoria Dias, Thomas Diehl, Katrina Dolendo, Franklin Dorschel, George Echeverria, Jordan Falk, Ethan Fan, Sayantika Ghosh, Liam Giombetti, Kelly Goonan, Aarushi Gupta, Akshay Gupta, Paula Gutierrez, Buchen (Olivia) Han, Olympia Hatzilambrou, Ryan Houser, Manya Jain, Rachael Johnson, Raynooka Kabir, Jaden Kielty, Grace Hyemin Kim, Hannah Laibinis, Ronit Langer, Angel Lee, Ga Ram Lee, Samuel Li, Jessica Lin, Catrina Malone, Lucca Maraston Oliveira, Megan McGuire, Meghan McQuillen, Kathryn Meadows, Brenna Means, Jayce Mei, Darius Meissner, Mackenzie Moore, Shoa Moosavi, Anusha Mudigonda, Misbah Nauman, Margaret Neely, Max Palys, Meera Parikh, Iktae Park, Luka Pauwelyn, Emily Pelles, Rachel Perkins, Ilona Ponyatyshyn, Sneha Puri, Pooja Reddy, Allie Reichert, Ryan Remmel, Emily Ren, Timothy Rudolph-Math, Neilah Rustemi, Helen Ryan, Beatrice Salas, Divya Sammeta, Grace Sander, Isabel Schaffer, Samantha Schlageter, Maclyn Senear, Kavya Shah, Emily Shambaugh, Emily Sherman, Kennedy Smith, Anna Strunjas, Alison Talty, May Tan, Joe Thomas, Krysten Thomas, Tyler Thompson, Briana Thrift, Zachary Trotzky, Allison Van Grinsven, Ileana Velez Alvarado, Danielle Venne, Sara Villanueva, Patrick Walsh, Jingxuan (Thomas) Wang, Yihao Wang, Sarah Weber, Ciara Weets, Courtney Wolf, Emily Woodrow, Theresa Worthington, Velen Wu, Wenhui Yang, Betelhem Yimer, Kayla Zamanian, Wei Zhang, Wenyu Zhu

## Competing interests

We have no conflicts of interest to report.

## Notes

### Competing Interest Statement

The authors have declared no competing interest.

### Summary of Updates

We have updated the manuscript to address reviewer comments. Specifically, the DOI has been updated to include a complete download of the data and corresponding files, we have updated discussion of other comparable datasets, and have replaced Figure 3 to highlight use cases for the data focused on the specific strengths of this dataset in comparison with others.

