## Supplementary Table 1 for "COVID AMP: An Open Access Dataset of COVID-19 Response Policies"

COVID ANALYSIS AND MAPPING OF POLICIES

### Supplementary Table 1: Data Dictionary

A description for each data column in the "Policies" tab and its possible values is provided below.

| | Unique ID | Authorizing entity | | | | |
|---|---|---|---|---|---|---|
| **Column name** | **Unique ID** | **Authorizing level of government** | **Authorizing country name** | **Authorizing country ISO** | **Authorizing state/province, if applicable** | **Authorizing local area (e.g., county, city)** |
| **Definition** | A unique identifier associated with data in each row.<br><br>The data is captured so that each row represents a single policy, per date issued, per authority and per area affected. | The level of government that authorized the policy | The name of the country in which the authorizing entity of the policy is located | The ISO-3 code of the authorizing country | The name of the sub-national area (e.g., state, province, district) corresponding to the country or agency which authorized the policy.<br><br>This value is "NA" if the policy was issued at the country level. | The name of the local area (e.g., city, county) corresponding to the country or agency which authorized the policy.<br><br>This value is "NA" if the policy was issued at the country or state/province level. |
| **Allowed values** | Numeric: Any unique integer value | One of:<br>Local<br>State / Province<br>Country | Text: Any country name (obtained from a pre-defined list based on ISO-3 code) | Text: Any 3-digit alpha-3 ISO code, as defined at https://www.iban.com/country-codes | Text: Any state, province, or other intermediate area name, or NA<br><br>Where possible, UN LOCODE geographic area names are used as defined at https://www.unece.org/cefact/codesfortrade/codes_index.html | Text: Any city, county, or other local area name, or NA |

| | | | | Affected area | |
|---|---|---|---|---|---|
| Authorizing local area (e.g., county, city) code | Authorizing role (e.g., official with authority) | Authorizing body (e.g., agency, office) | Name of official, if relevant | Affected level of government | Affected country name |
| If available, a unique ID for the local area (e.g., city, county) corresponding to the location which authorized the policy.<br><br>This value is "N/A" if the policy was issued at the country or state/province level, or "Undefined" if a unique ID is not defined in the AMP dataset. | The name of the entity who authorized the policy.<br><br>This value is "NA" if the information was not available or was not relevant (e.g., in the case of judicial rulings). | The office of the authorizing entity that issued the policy | If relevant, the first and last name of the authorizing official | The level of government to which the policy applies.<br><br>This information is only included if the level of government authorizing a policy - see "Authorizing level of government" - is different from that affected (e.g., a state authorizes a policy that affects specific counties). | The name of the country to which the policy applies |
| Text: Any unique ID, such as a 5-digit FIPS code for USA counties, or NA or Undefined | Text: Any role | Text: Any office name, agency, or similar entity name | Text: Any name | One of:<br>Local<br>State / Province<br>Country | Text: Any country name (obtained from a pre-defined list based on ISO-3 code) |

| | | | | Policy/Law |
|---|---|---|---|---|
| **Affected country ISO** | **Affected state/province (intermediate area)** | **Affected local area (e.g., county, city)** | **Affected local area (e.g., county, city) code** | **Policy relaxing or restricting** |
| The ISO-3 code of the affected country | The name of the sub-national area (e.g., state, province, district) affected by the policy.<br><br>This value is "NA" if the affected location is the same as the authorizing location (e.g., a state authorizing policy that only affects that state). | The name of the local area (e.g., city, county) affected by the policy/law.<br><br>This value is "NA" if the affected location is the same as the authorizing location (e.g., a city authorizing policy that only affects that city). | If available, a unique ID for the local area (e.g., city, county) affected by the policy.<br><br>This value is "N/A" if the policy was issued at the country or state/province level, or "Undefined" if a unique ID is not defined in the AMP dataset. | Broad designations about a policy with regard to its intended impact on the policy environment at the time the policy was issued. |
| Text: Any 3-digit alpha-3 ISO code, as defined at https://www.iban.com/country-codes | Text: Any state, province, or other intermediate area name, or NA<br><br>Where possible, UN LOCODE geographic area names are used as defined at https://www.unece.org/cefact/codesfortrade/codes_index.html | Text: Any city, county, or other local area name, or NA | Text: Any unique ID, such as a 5-digit FIPS code for USA counties, or NA or Undefined | One of:<br>Relaxing<br>Restricting<br>Other |

| Policy category | Policy subcategory | Policy target | Policy description | Issued date |
|---|---|---|---|---|
| Categorization of the overall scope of a policy based on its content | Detailed information about the intention of the policy based on its content | The primary population, location or entities impacted by the policy or law | A written description of the policy and the directive by the researcher | The date on which the policy or law was initially announced and/or issued |
| One of:<br>Authorizing and enforcement<br>Contact tracing/Testing<br>Emergency declarations<br>Enabling and relief measures<br>Face mask<br>Military mobilization<br>Social distancing<br>Support for public health and clinical capacity<br>Travel restrictions<br>Vaccinations | Any of:<br>Authorization; Enforcement; Contact tracing; Testing; General emergency declaration; Public health emergency declaration; Anti-price gouging measures; Budget modifications; Early prison release; Eviction and foreclosure delays; Extension of public services; Hazard pay; Leave entitlement adjustments; Modification of unemployment benefits; Mortgage payment support; Other labor protections; Other relief measures; Regulatory relief; Relief funding; Remote notarization; Stimulus payments; Support for essential workers; Tax delay; Utility payment; Face mask required; Face mask suggested; Face mask exemption; Face mask (other); Activation of military for enforcement; Activation of military for logistical and/or medical support; Adaptation and mitigation measures; Alternative election measures; Curfews; Distancing mandate; Event delays or cancellations; Face covering; Health screening; Isolation; Lockdown; Mass gathering restrictions; Other forms of social distancing; Quarantine; Prison population reduction; Private sector closures; Public service closures; Safer at home; School closures; Stay at home; Visitor restrictions; Coverage for cost of testing; Crisis standards of care; Elective procedure delay or cancellation; Emergency use or expanded market authorization; Healthcare facility licensing waivers; Immunity for medical providers; Medical licensing waivers; Notification requirements; Other measures to support public health and clinical capacity; Revised "emergency personnel" designations; Risk communication; Support for telemedicine; Domestic travel restriction; Domestic travel restrictions (interstate); Domestic travel restrictions (intrastate); International travel restriction; Multi-vaccine policy; Vaccine administration, distribution, and logistics; Vaccine cost, financing, and insurance; Vaccine exemption or alternative; Vaccine mandate; Vaccine prioritization; Vaccine-related plan | Any of:<br>All essential businesses; All non essential businesses; All recreation/entertainment venues; All workers/employees; Bars/alcohol service establishments; Businesses/private sector; Casinos/gambling facilities; Childcare facilities (preschool or other daycare); Churches/places of worship; Confirmed cases; Courthouse/judiciary; Criminal justice system/prisons/jails/incarcerated persons; Cultural events/weddings/funerals; Day camps/summer camps; Essential workers; Event center/convention/performance hall/fair; Family entertainment/leisure activity; Farmer's market/open air market; Farming/agriculture/food processing; Fully vaccinated individuals; Funeral home/mortuary; General population; Grocery store; Gym/fitness center/recreational sports facility; Healthcare workers and allied personnel; Higher education; Homeless shelters/homeless individuals; Hospitals/urgent care/emergency medical services; Hotels/lodging; Housing sector/real estate/renters/homeowners; Indigenous people; Insurance/banking; Libraries/museums/art galleries/community centers; Liquor stores; Manufacturers; Marginalized communities; Medical offices (doctor/dentist/physical therapist); Migrant workers/refugees/asylum seekers; Military; Minors and children; Night club/dance hall/karaoke; Non-essential workers; Non-governmental organizations; Notary; Nursing homes/assisted living facility; Older adults/individuals with underlying medical conditions; Omicron; Organized adult sports; Outdoor recreation/campgrounds/beaches/parks; Personal care facilities; Pharmacy; Primary education; Public administration; Public sector/public buildings/government employees; Public services; Public transportation; Restaurants/food service establishments; Retail stores; Secondary education; Small businesses; Stadiums/arenas/racetracks/sporting venues; Suspected cases; Swimming pools; Theatres/movie theatres; Transportation sector/shipping/logistics; Travelers; Unemployed/underemployed workers; University/professional sports; Unvaccinated/partially vaccinated individuals; Veterinary clinics; Youth/school sports/extracurricular activities | Text: Any English-language text | Date: format mm/dd/yyyy |

| Effective start date | Anticipated end date | Actual end date | Intended duration | Prior row ID linked to this entry | Data source for policy announcement |
|---|---|---|---|---|---|
| The date on which the policy took effect or was enacted | The date on which the directive specified in the policy was intended to end.<br><br>If a specific measure was extended beyond the initial end date, each extension is captured as a new row. Extended policies are linked back to the previous policy through the field 'Prior row ID linked to this entry'. | The date on which the directive specified in the policy was terminated, replaced, or extended. | A text description of the intended duration of the policy, expressed in days where applicable | The unique ID of the prior policy that immediately preceded the specified policy.<br><br>This information is included when a policy provided an update or extension of a prior policy already captured by the dataset or modifies the original policy in some way. This is intended to capture policies that are extended past their initial anticipated end date. In all other cases, this field will be blank. | A link to the website from which the policy data were gathered. This link may be a news report or press announcement if data were gathered from that source and not from the source policy document. |
| Date: format mm/dd/yyyy | Date: Any date after the specified effective start date, format mm/dd/yyyy | Date: Any date after the specified effective start date, format mm/dd/yyyy | Text: Any text value | Numeric: Blank, or any unique policy ID | Any URL |

| Policy/law name | Policy/law type | Data source for law/policy | PDF file name of law/policy | Attachment for policy | Policy number |
|---|---|---|---|---|---|
| The complete title of the law, policy, or policy announcement, including any relevant numerical information. | The type of law or policy that is enacted | Source documentation for the underlying law or policy | The file name of the PDF file of the corresponding law or policy | URL of permanently hosted PDF document(s) for the policy | Policy number is a numeric identifier given to each policy release, including capturing co-released policies, where applicable. A single policy release may contain multiple directives. For example, an executive order can include a stay at home order for individuals and mandate non-essential business closures. These policy elements will share a single Policy Number since they are part of a larger order/law/legislation, but they will be captured in separate lines with different Unique IDs. |
| Text: Any text value | One of:<br>Case<br>Declaration<br>Directive<br>Mandate<br>Memorandum<br>Non-policy guidance<br>Order<br>Ordinance<br>Proclamation<br>Regulation<br>Statute | Any URL | Text: Any file name (PDF) | Any URL(s) | Numeric: A numeric identifier created from the PDF file name and the Policy/law name |

#### Legal authority

| Authorizing entity has authority? | Relevant authority (e.g., statute) to make the law/policy | Data source for authority to make the law/policy | Home rule state? | Dillon's Rule State |
|---|---|---|---|---|
| Whether the entity who authorized the policy has the legal authority to do so, at the time that the policy was issued or effective.<br><br>When a policy is under legal challenge, the field "Legal challenge?" will be set to Yes. | Title of legal authority for entity to enact the policy | Source documentation for the relevant authority specified | Whether the state is a "Home Rule" state or a "Limited Home Rule" state | Whether or not a state is under Dillon's rule. |
| One of:<br>Yes<br>No<br>Unclear | Text: Any text value | Any URL | One of:<br>Yes<br>Limited<br>No | One of:<br>Yes<br>Limited<br>No<br>Unclear |
