## Supplementary Table 2 for "COVID AMP: An Open Access Dataset of COVID-19 Response Policies"

COVID ANALYSIS AND MAPPING OF POLICIES

### Supplementary Table 2: Policy count by country

| Country name | Policy count |
|---|---|
| United States of America (USA) | 28634 |
| Canada (CAN) | 2558 |
| France (FRA) | 1334 |
| Denmark (DNK) | 771 |
| Brazil (BRA) | 759 |
| Spain (ESP) | 653 |
| Czechia (CZE) | 607 |
| South Africa (ZAF) | 599 |
| Philippines (PHL) | 490 |
| Hungary (HUN) | 431 |
| Belgium (BEL) | 413 |
| Bolivia (Plurinational State of) (BOL) | 388 |
| Russian Federation (RUS) | 376 |
| Estonia (EST) | 369 |
| India (IND) | 362 |
| Korea, Republic of (KOR) | 354 |
| Thailand (THA) | 353 |
| Argentina (ARG) | 352 |
| England (GB-ENG) | 343 |
| Scotland (GB-SCT) | 318 |
| Germany (DEU) | 318 |
| Peru (PER) | 268 |
| Norway (NOR) | 266 |
| Panama (PAN) | 259 |
| Italy (ITA) | 258 |
| Serbia (SRB) | 243 |
| New Zealand (NZL) | 239 |
| Sweden (SWE) | 209 |
| Botswana (BWA) | 145 |
| Australia (AUS) | 141 |
| Colombia (COL) | 141 |
| Chile (CHL) | 124 |
| Chad (TCD) | 117 |
| Fiji (FJI) | 117 |
| Hong Kong (HKG) | 116 |
| Nigeria (NGA) | 114 |
| Uruguay (URY) | 111 |
| Egypt (EGY) | 106 |
| Ireland (IRL) | 105 |
| China (CHN) | 100 |
| Japan (JPN) | 89 |
| Uganda (UGA) | 87 |
| Saudi Arabia (SAU) | 86 |
| Mexico (MEX) | 83 |
| Iraq (IRQ) | 83 |
| Oman (OMN) | 82 |
| Ghana (GHA) | 81 |
| Tunisia (TUN) | 79 |
| Congo, Democratic Republic of the (COD) | 74 |
| El Salvador (SLV) | 74 |
| Marshall Islands (MHL) | 73 |
| Djibouti (DJI) | 72 |
| Northern Ireland (GB-NIR) | 70 |
| Ecuador (ECU) | 69 |
| Niger (NER) | 69 |
| Taiwan, Province of China (TWN) | 67 |
| Singapore (SGP) | 64 |
| Morocco (MAR) | 64 |
| Senegal (SEN) | 63 |
| Libya (LBY) | 63 |
| Tanzania, United Republic of (TZA) | 59 |
| Jordan (JOR) | 59 |
| Côte d'Ivoire (CIV) | 58 |
| Wales (GB-WLS) | 58 |
| Israel (ISR) | 55 |
| Lebanon (LBN) | 45 |
| Switzerland (CHE) | 41 |
| Yemen (YEM) | 28 |
| Ethiopia (ETH) | 22 |

| | |
|---|---|
| Honduras (HND) | 17 |
| Mongolia (MNG) | 12 |
| Burkina Faso (BFA) | 10 |
| Kenya (KEN) | 7 |
| Zimbabwe (ZWE) | 5 |
| Bahrain (BHR) | 4 |
| Finland (FIN) | 4 |
| Rwanda (RWA) | 4 |
| Ukraine (UKR) | 3 |
| Indonesia (IDN) | 3 |
| Qatar (QAT) | 3 |
| Sudan (SDN) | 3 |
| Pakistan (PAK) | 3 |
| Croatia (HRV) | 3 |
| Zambia (ZMB) | 3 |
| Gambia (GMB) | 2 |
| United Arab Emirates (ARE) | 2 |
| Eswatini (SWZ) | 2 |
| Sri Lanka (LKA) | 2 |
| Uzbekistan (UZB) | 2 |
| Cyprus (CYP) | 2 |
| Mozambique (MOZ) | 2 |
| Seychelles (SYC) | 2 |
| Cambodia (KHM) | 2 |
| Mauritius (MUS) | 2 |
| Liberia (LBR) | 2 |
| Romania (ROU) | 2 |
| Latvia (LVA) | 2 |
| Curaçao (CUW) | 2 |
| Luxembourg (LUX) | 2 |
| Aruba (ABW) | 2 |
| Guatemala (GTM) | 2 |
| Malaysia (MYS) | 2 |
| Somalia (SOM) | 2 |
| Bahamas (BHS) | 2 |
| Portugal (PRT) | 2 |
| Turkey (TUR) | 1 |
| Armenia (ARM) | 1 |
| Malta (MLT) | 1 |
| Saint Kitts and Nevis (KNA) | 1 |
| Togo (TGO) | 1 |
| Saint Vincent and the Grenadines (VCT) | 1 |
| American Samoa (ASM) | 1 |
| Cuba (CUB) | 1 |
| Albania (ALB) | 1 |
| Samoa (WSM) | 1 |
| Palau (PLW) | 1 |
| Myanmar (MMR) | 1 |
| Trinidad and Tobago (TTO) | 1 |
| Dominican Republic (DOM) | 1 |
| Maldives (MDV) | 1 |
| Austria (AUT) | 1 |
| Poland (POL) | 1 |
| Gabon (GAB) | 1 |
| Antigua and Barbuda (ATG) | 1 |
| Nepal (NPL) | 1 |
| Cayman Islands (CYM) | 1 |
| Netherlands (NLD) | 1 |
| Bangladesh (BGD) | 1 |
| Greece (GRC) | 1 |
| Sint Maarten (Dutch part) (SXM) | 1 |
| Iran (Islamic Republic of) (IRN) | 1 |
| Slovakia (SVK) | 1 |
| Paraguay (PRY) | 1 |
| Slovenia (SVN) | 1 |
| Tuvalu (TUV) | 1 |
| Jamaica (JAM) | 1 |
| European Union (EU) | 1 |
| Georgia (GEO) | 1 |
| Angola (AGO) | 1 |
| Brunei Darussalam (BRN) | 1 |
| Puerto Rico (PRI) | 1 |
| Bulgaria (BGR) | 1 |
| Lithuania (LTU) | 1 |
| Iceland (ISL) | 1 |
| Kuwait (KWT) | 1 |
| Moldova, Republic of (MDA) | 1 |
| Suriname (SUR) | 1 |

| | |
|---|---|
| Lesotho (LSO) | 1 |
| Macao (MAC) | 1 |
| Malawi (MWI) | 1 |

COVID ANALYSIS AND MAPPING OF POLICIES

#### Supplementary Table 2: Policy count by intermediate and local areas

| Jurisdiction name | Policy count |
|---|---|
| **United States of America (USA)** | **28634** |
| **Alabama** | **152** |
| Jefferson County, AL | 6 |
| Montgomery (city), AL | 1 |
| **Alaska** | **150** |
| Juneau City and Borough, AK | 6 |
| **Arizona** | **249** |
| Coconino County, AZ | 5 |
| Kingman (city), AZ | 4 |
| Lake Havasu City (city), AZ | 4 |
| Maricopa County, AZ | 2 |
| Mohave County, AZ | 3 |
| Phoenix (city), AZ | 5 |
| Pima County, AZ | 14 |
| Pinal County, AZ | 1 |
| Tucson (city), AZ | 44 |
| **Arkansas** | **145** |
| Little Rock (city), AR | 6 |
| **California** | **3855** |
| Alameda County, CA | 116 |
| Alpine County, CA | 20 |
| Amador County, CA | 11 |
| Berkeley (city), CA | 8 |
| Butte County, CA | 15 |
| Calaveras County, CA | 30 |
| Colusa County, CA | 5 |
| Contra Costa County, CA | 120 |
| Del Norte County, CA | 25 |
| El Dorado County, CA | 16 |
| Fresno (city), CA | 156 |
| Fresno County, CA | 215 |
| Glenn County, CA | 35 |
| Humboldt County, CA | 21 |
| Imperial County, CA | 101 |
| Inyo County, CA | 32 |
| Kern County, CA | 12 |
| Kings County, CA | 7 |
| Lake County, CA | 29 |
| Lassen County, CA | 6 |
| Los Angeles (city), CA | 153 |
| Los Angeles County, CA | 69 |
| Madera County, CA | 36 |
| Marin County, CA | 108 |
| Mariposa County, CA | 116 |
| Mendocino County, CA | 436 |
| Merced County, CA | 78 |
| Mono County, CA | 44 |
| Monterey County, CA | 31 |
| Napa County, CA | 8 |
| Nevada County, CA | 6 |
| Oakland (city), CA | 21 |
| Orange County, CA | 242 |
| Placer County, CA | 9 |
| Plumas County, CA | 18 |
| Red Bluff (city), CA | 1 |
| Redwood City (city), CA | 1 |
| Riverside County, CA | 18 |
| Sacramento (city), CA | 4 |
| Sacramento County, CA | 51 |
| San Benito County, CA | 5 |
| San Bernardino (city), CA | 3 |
| San Bernardino County, CA | 21 |
| San Diego (city), CA | 41 |
| San Diego County, CA | 169 |
| San Francisco (city), CA | 115 |
| San Joaquin County, CA | 37 |
| San Jose (city), CA | 1 |
| San Luis Obispo County, CA | 23 |

| | |
|---|---|
| San Mateo County, CA | 61 |
| Santa Barbara County, CA | 100 |
| Santa Clara County, CA | 20 |
| Santa Cruz County, CA | 69 |
| Shasta County, CA | 4 |
| Sierra County, CA | 7 |
| Siskiyou County, CA | 6 |
| Solano County, CA | 25 |
| Sonoma County, CA | 57 |
| Stanislaus County, CA | 28 |
| Sutter County, CA | 38 |
| Trinity County, CA | 5 |
| Tulare County, CA | 5 |
| Tuolumne County, CA | 62 |
| Ventura County, CA | 48 |
| Yolo County, CA | 23 |
| Yuba County, CA | 38 |
| **Colorado** | **756** |
| Denver County, CO | 5 |
| **Connecticut** | **454** |
| **Delaware** | **231** |
| Dover (city), DE | 1 |
| **District of Columbia** | **526** |
| **Florida** | **692** |
| Alachua County, FL | 80 |
| Broward County, FL | 106 |
| Deland (city), FL | 1 |
| Hillsborough County, FL | 1 |
| Jacksonville (city), FL | 1 |
| Leon County, FL | 1 |
| Martin County, FL | 1 |
| Miami-Dade County, FL | 201 |
| Orange County, FL | 1 |
| Palm Beach County, FL | 76 |
| Pinellas County, FL | 1 |
| Seminole County, FL | 2 |
| St. Augustine (city), FL | 1 |
| Tallahassee (city), FL | 3 |
| Walton County, FL | 2 |
| **Georgia** | **921** |
| Atlanta (city), GA | 17 |
| Cherokee County, GA | 20 |
| Clayton County, GA | 16 |
| Cobb County, GA | 4 |
| DeKalb County, GA | 51 |
| Fulton County, GA | 9 |
| Gordon County, GA | 9 |
| Gwinnett County, GA | 17 |
| **Guam** | **10** |
| **Hawaii** | **947** |
| Hawai'i County | 24 |
| Honolulu County, HI | 249 |
| Kaua'i County | 26 |
| Maui County | 165 |
| **Idaho** | **206** |
| Ada County, ID | 36 |
| Boise City (city), ID | 65 |
| **Illinois** | **519** |
| Chicago (city), IL | 83 |
| Cook County, IL | 27 |
| Springfield (city), IL | 4 |
| **Indiana** | **243** |
| **Iowa** | **329** |
| **Kansas** | **260** |
| Clay County, KS | 1 |
| Douglas County, KS | 22 |
| Johnson County, KS | 15 |
| Leavenworth County, KS | 6 |
| Wyandotte County, KS | 24 |
| **Kentucky** | **179** |
| Louisville (city), KY | 1 |
| **Louisiana** | **661** |
| **Maine** | **167** |
| **Maryland** | **852** |
| Baltimore City, MD | 140 |
| Bowie (city), MD | 1 |
| Charles County, MD | 7 |
| College Park (city), MD | 1 |

| | |
|---|---|
| Frederick (city), MD | 20 |
| Frederick County, MD | 10 |
| Gaithersburg (city), MD | 4 |
| Greenbelt (city), MD | 1 |
| Hyattsville (city), MD | 5 |
| Laurel (city), MD | 134 |
| Montgomery County, MD | 70 |
| Prince George's County, MD | 74 |
| Rockville (city), MD | 17 |
| Takoma Park (city), MD | 16 |
| **Massachusetts** | **471** |
| Boston (city), MA | 95 |
| Braintree Town (city), MA | 2 |
| Cambridge (city), MA | 45 |
| Suffolk County, MA | 1 |
| Worcester County, MA | 1 |
| **Michigan** | **187** |
| **Minnesota** | **303** |
| **Mississippi** | **510** |
| Adams County, MS | 1 |
| Alcorn County, MS | 2 |
| Benton County, MS | 1 |
| Coahoma County, MS | 1 |
| DeSoto County, MS | 1 |
| Lafayette County, MS | 1 |
| Marshall County, MS | 1 |
| Oxford (city), MS | 2 |
| Panola County, MS | 1 |
| Quitman County, MS | 1 |
| Tippah County, MS | 1 |
| Tunica County, MS | 1 |
| **Missouri** | **183** |
| Kansas City (city), MO | 58 |
| St. Louis County, MO | 1 |
| **Montana** | **89** |
| **Nebraska** | **876** |
| **Nevada** | **412** |
| Carson City, NV | 5 |
| Churchill County, NV | 12 |
| Clark County, NV | 18 |
| Douglas County, NV | 5 |
| Elko County, NV | 16 |
| Esmeralda County, NV | 11 |
| Eureka County, NV | 22 |
| Humboldt County, NV | 9 |
| Lander County, NV | 10 |
| Lincoln County, NV | 8 |
| Lyon County, NV | 9 |
| Mineral County, NV | 3 |
| Nye County, NV | 27 |
| Pershing County, NV | 7 |
| Storey County, NV | 5 |
| Washoe County, NV | 13 |
| White Pine County, NV | 16 |
| **New Hampshire** | **665** |
| **New Jersey** | **759** |
| **New Mexico** | **524** |
| **New York** | **1512** |
| Albany (city), NY | 20 |
| New York City (city), NY | 586 |
| **North Carolina** | **365** |
| **North Dakota** | **115** |
| **Ohio** | **352** |
| **Oklahoma** | **569** |
| **Oregon** | **365** |
| **Pennsylvania** | **283** |
| **Puerto Rico** | **341** |
| **Rhode Island** | **919** |
| **South Carolina** | **319** |
| Columbia (city), SC | 41 |
| **South Dakota** | **158** |
| Sioux Falls, SD | 6 |
| **Tennessee** | **573** |
| Davidson County, TN | 290 |
| **Texas** | **1664** |
| Anderson County, TX | 2 |
| Andrews County, TX | 7 |
| Bexar County, TX | 174 |

| | |
|---|---|
| Dallas County, TX | 427 |
| El Paso (city), TX | 283 |
| El Paso County, TX | 149 |
| Tarrant County, TX | 60 |
| Travis County, TX | 67 |
| **Utah** | **738** |
| **Vermont** | **508** |
| **Virginia** | **491** |
| Alexandria City, VA | 35 |
| Arlington County, VA | 39 |
| Fairfax City, VA | 16 |
| Fairfax County, VA | 29 |
| Falls Church City, VA | 2 |
| Fauquier County, VA | 1 |
| Loudoun County, VA | 30 |
| Manassas City, VA | 16 |
| Manassas Park City, VA | 22 |
| Prince William County, VA | 38 |
| Warren County, VA | 18 |
| **Washington** | **989** |
| King County, WA | 5 |
| Seattle (City), WA | 37 |
| **West Virginia** | **157** |
| **Wisconsin** | **145** |
| **Wyoming** | **470** |
| **Canada (CAN)** | **2558** |
| **Alberta** | **126** |
| **British Columbia** | **326** |
| **Manitoba** | **17** |
| **N/A** | **409** |
| **New Brunswick** | **277** |
| **Northwest Territories** | **164** |
| **Nova Scotia** | **84** |
| **Nunavut** | **55** |
| **Ontario** | **82** |
| **Quebec** | **343** |
| **Saskatchewan** | **570** |
| **Yukon** | **105** |
| **France (FRA)** | **1334** |
| **Île-de-France** | **2** |
| **Denmark (DNK)** | **771** |
| **Brazil (BRA)** | **759** |
| **São Paulo** | **130** |
| **Spain (ESP)** | **653** |
| **Czechia (CZE)** | **607** |
| **South Africa (ZAF)** | **599** |
| **Philippines (PHL)** | **490** |
| **Hungary (HUN)** | **431** |
| **Belgium (BEL)** | **413** |
| **Bruxelles-Capitale** | **58** |
| **Communauté Française** | **3** |
| **Flanders** | **16** |
| **Liège/Luik** | **32** |
| **Wallonie** | **11** |
| **Bolivia (Plurinational State of) (BOL)** | **388** |
| **Santa Cruz** | **111** |
| Santa Cruz de la Sierra | 111 |
| **Russian Federation (RUS)** | **376** |
| **Moscow Oblast** | **263** |
| Moscow (City), Moscow Oblast | 263 |
| **Estonia (EST)** | **369** |
| **India (IND)** | **362** |
| **Goa** | **3** |
| North Goa, Goa | 2 |
| South Goa, Goa | 1 |
| **Maharashtra** | **4** |
| **National Capital Territory of Delhi (union te** | **5** |
| Delhi (union territory) | 4 |
| **Tamil Nadu** | **2** |
| **Korea, Republic of (KOR)** | **354** |
| **Busan City** | **1** |
| Busan City | 1 |
| **Seoul Special City** | **20** |
| Seoul Special City | 20 |
| **Thailand (THA)** | **353** |
| **Krung Thep Maha Nakhon (Bangkok)** | **4** |
| **Argentina (ARG)** | **352** |
| **England (GB-ENG)** | **343** |

| | |
|---|---|
| **Scotland (GB-SCT)** | **318** |
| **Germany (DEU)** | **318** |
| **White Mountain Apache Reservation** | **274** |
| **Peru (PER)** | **268** |
| **Lima Province** | **9** |
| Ate, Lima Province | 1 |
| La Molina, Lima Province | 2 |
| Lima Norte, Lima Province | 1 |
| Puente Piedra, Lima Province | 1 |
| San Isidro, Lima Province | 1 |
| San Martín de Porres, Lima Province | 3 |
| **Norway (NOR)** | **266** |
| **Panama (PAN)** | **259** |
| **Italy (ITA)** | **258** |
| **Lombardia** | **1** |
| **Veneto** | **1** |
| **Serbia (SRB)** | **243** |
| **New Zealand (NZL)** | **239** |
| **Sweden (SWE)** | **209** |
| **Navajo Nation** | **192** |
| **Blackfeet Nation** | **191** |
| **Botswana (BWA)** | **145** |
| **Australia (AUS)** | **141** |
| **Australian Capital Territory** | **10** |
| **New South Wales** | **10** |
| **Northern Territory** | **14** |
| **Queensland** | **15** |
| **South Australia** | **14** |
| **Tasmania** | **13** |
| **Victoria** | **25** |
| **Western Australia** | **19** |
| **Colombia (COL)** | **141** |
| **Colorado River Indian Tribes** | **127** |
| **Chile (CHL)** | **124** |
| **Chad (TCD)** | **117** |
| **Fiji (FJI)** | **117** |
| **Hong Kong (HKG)** | **116** |
| **Nigeria (NGA)** | **114** |
| **Fort McDowell Yavapai Nation** | **111** |
| **Uruguay (URY)** | **111** |
| **Gila River Indian Community** | **110** |
| **Egypt (EGY)** | **106** |
| **Ireland (IRL)** | **105** |
| **China (CHN)** | **100** |
| **Japan (JPN)** | **89** |
| **Tokyo Metropolis** | **1** |
| **Oglala Sioux Tribe** | **88** |
| **Uganda (UGA)** | **87** |
| **Saudi Arabia (SAU)** | **86** |
| **Najran** | **1** |
| **Mexico (MEX)** | **83** |
| **Iraq (IRQ)** | **83** |
| **Oman (OMN)** | **82** |
| **Ghana (GHA)** | **81** |
| **Tunisia (TUN)** | **79** |
| **Prairie Band Potawatomi Nation** | **75** |
| **Congo, Democratic Republic of the (COD)** | **74** |
| **El Salvador (SLV)** | **74** |
| **Marshall Islands (MHL)** | **73** |
| **Djibouti (DJI)** | **72** |
| **Northern Ireland (GB-NIR)** | **70** |
| **Ecuador (ECU)** | **69** |
| **Niger (NER)** | **69** |
| **Taiwan, Province of China (TWN)** | **67** |
| **Singapore (SGP)** | **64** |
| **Morocco (MAR)** | **64** |
| **Marrakesh-Safi** | **3** |
| Ouezzane (Wazan) Province | 1 |
| **Senegal (SEN)** | **63** |
| **Libya (LBY)** | **63** |
| **Sac and Fox Nation** | **61** |
| **Tanzania, United Republic of (TZA)** | **59** |
| **Wales (GB-WLS)** | **58** |
| **Lebanon (LBN)** | **45** |
| **Tohono O'odham** | **42** |
| **Switzerland (CHE)** | **41** |
| **Hualapai Tribe** | **31** |
| **Yemen (YEM)** | **28** |

| | |
|---|---|
| Crow Nation | 22 |
| Ethiopia (ETH) | 22 |
| Hopi Tribe | 19 |
| Honduras (HND) | 17 |
| Mongolia (MNG) | 12 |
| Fort Yuma Quechan Indian Tribe | 11 |
| Shoshone-Paiute Tribe | 11 |
| Salt River Pima-Maricopa Indian Community | 11 |
| Reno-Sparks Indian Colony | 11 |
| Walker River Paiute Tribe | 10 |
| Burkina Faso (BFA) | 10 |
| Washoe Tribe | 9 |
| Fort Mojave Tribe | 9 |
| Kenya (KEN) | 7 |
| Ak-Chin Indian Community | 7 |
| Yerington Paiute Tribe | 7 |
| Zimbabwe (ZWE) | 5 |
| Bahrain (BHR) | 4 |
| Finland (FIN) | 4 |
| Rwanda (RWA) | 4 |
| Pyramid Lake Paiute Tribe | 3 |
| Ukraine (UKR) | 3 |
| Indonesia (IDN) | 3 |
| Qatar (QAT) | 3 |
| Iowa Tribe of Kansas and Nebraska | 3 |
| Sudan (SDN) | 3 |
| Pakistan (PAK) | 3 |
| Croatia (HRV) | 3 |
| Zambia (ZMB) | 3 |
| Gambia (GMB) | 2 |
| United Arab Emirates (ARE) | 2 |
| Eswatini (SWZ) | 2 |
| Sri Lanka (LKA) | 2 |
| Uzbekistan (UZB) | 2 |
| Cyprus (CYP) | 2 |
| Mozambique (MOZ) | 2 |
| Summit Lake Paiute Tribe | 2 |
| Seychelles (SYC) | 2 |
| Cambodia (KHM) | 2 |
| Mauritius (MUS) | 2 |
| Liberia (LBR) | 2 |
| Romania (ROU) | 2 |
| Latvia (LVA) | 2 |
| Curaçao (CUW) | 2 |
| Luxembourg (LUX) | 2 |
| Aruba (ABW) | 2 |
| Guatemala (GTM) | 2 |
| Malaysia (MYS) | 2 |
| Somalia (SOM) | 2 |
| Bahamas (BHS) | 2 |
| Portugal (PRT) | 2 |
| Turkey (TUR) | 1 |
| Armenia (ARM) | 1 |
| Malta (MLT) | 1 |
| Saint Kitts and Nevis (KNA) | 1 |
| Togo (TGO) | 1 |
| Saint Vincent and the Grenadines (VCT) | 1 |
| American Samoa (ASM) | 1 |
| Cuba (CUB) | 1 |
| Albania (ALB) | 1 |
| Samoa (WSM) | 1 |
| Palau (PLW) | 1 |
| Myanmar (MMR) | 1 |
| Trinidad and Tobago (TTO) | 1 |
| Dominican Republic (DOM) | 1 |
| Maldives (MDV) | 1 |
| Austria (AUT) | 1 |
| Poland (POL) | 1 |
| Gabon (GAB) | 1 |
| Antigua and Barbuda (ATG) | 1 |
| Nepal (NPL) | 1 |
| Cayman Islands (CYM) | 1 |
| Netherlands (NLD) | 1 |
| Bangladesh (BGD) | 1 |
| Duckwater Shoshone Tribe | 1 |
| Greece (GRC) | 1 |
| Sint Maarten (Dutch part) (SXM) | 1 |

| | |
|---|---|
| Iran (Islamic Republic of) (IRN) | 1 |
| Slovakia (SVK) | 1 |
| Paraguay (PRY) | 1 |
| Slovenia (SVN) | 1 |
| Tuvalu (TUV) | 1 |
| Jamaica (JAM) | 1 |
| European Union (EU) | 1 |
| Georgia (GEO) | 1 |
| Angola (AGO) | 1 |
| Brunei Darussalam (BRN) | 1 |
| Big Pine Paiute Tribe of the Owens Valley | 1 |
| Bulgaria (BGR) | 1 |
| Lithuania (LTU) | 1 |
| Iceland (ISL) | 1 |
| Kuwait (KWT) | 1 |
| Moldova, Republic of (MDA) | 1 |
| Suriname (SUR) | 1 |
| Lesotho (LSO) | 1 |
| Macao (MAC) | 1 |
| Malawi (MWI) | 1 |
